# Factors associated with pharmacologic serum concentration of Magnesium and corresponding obstetric outcomes among women with severe pre-eclampsia at Iringa Regional Referral Hospital

**DOI:** 10.1101/2024.08.21.24312387

**Authors:** Joshua Stephen Kileo, Shubi matovelo, Gabriel Kitinusa

## Abstract

**Introduction:** Severe pre-eclampsia is managed by using Magnesium sulphate to prevent its progress into eclampsia with a success rate of more than 50%. This study aimed at establishing reliable data pertaining factors influencing pharmacologic serum concentration of Magnesium and their correlation to obstetric outcomes.

**Methods:** This study was carried out at Iringa regional referral hospital, spanning six months from November 2023 to April 2024. A convenient sampling technique was used to recruit 121 pre-eclamptic patients. Recruited women were tested serum Magnesium at admission, 4 and 12 hours post initial administration of MgSO_4_. IBM SPSS version 29 was used for analysis where, Continuous variables were analyzed by using median and interquartile range while categorical variables were analyzed using frequency and percentages. The associations between variables were determined using chi-square tests, univariate, and multivariate binary regression models. A p value<0.05 was considered statistically significant.

**Results:** The median age of participants were (29±10 IQR), the median serum Mg at admission was (0.8±0.29 IQR) Mmol/L majority with low 53(43.8%) levels. Proportion of subtherapeutic serum Mg was 38.8% with median of (1.9±1.02 IQR) Mmol/L. Multivariate regression showed that; low serum Mg at admission [p<0.001, OR= 9.17, 95% CI (3.07-27.37)], Creatinine level [p = 0.012, OR= 4.49, 95% CI (1.38-14.6)], Overweight [p= 0.002, OR= 5.52, 95% CI (1.83-16.68)] and proteinuria +++[p-value = 0.004, OR= 7.13, 95% CI (1.84-26.86)] were significantly associated with subtherapeutic level. After adjusting with other factors, it was found that subtherapeutic level [p<0.001, OR=25.44(5.73-112.90)], elevated creatinine [p=0.040, OR=3.59(1.66-12.15)], Reduced urine output [p=0.001, OR=6.69(2.15-20.90)] were significant predictors of adverse maternal outcomes.

**Conclusion:** Subtherapeutic serum Magnesium which significantly predict adverse obstetric outcomes, is influenced by factors which can be monitored and be useful in effective management of severe pre-eclampsia. This can help in administering tailored dose of MgSO_4_ for better outcomes.

## Introduction

Severe pre-eclampsia is among major pregnancy related complications characterized by high blood pressure and potential damage to organs. It is the third most common cause of maternal and fetal mortality globally only after severe bleeding i.e., postpartum hemorrhage and infections (Kreepala, Kitporntheranunt, Rungsrithananon, & Wattanavaekin, 2018; Mirkovic, Tulic, Stankovic, & Soldatovic, 2020; Padda et al., 2021a; Shepherd et al., 2019).

The prevalence of severe pre-eclampsia in developing countries is high, estimated to range from 4% to 18% (Altraigey and Mostafaa 2019; Edward et al. 2021; Kreepala et al. 2018). Severe pre-eclampsia causes a substantial health risk mostly in low- and middle-income countries because of challenges related to limited access to adequate antenatal care services. Severe pre-eclampsia causes maternal and fetal mortality because management of women with complications of severe pre-eclampsia like organ failure is challenging and management of premature babies being the challenge as well (Darkwa et al., 2017; USAID, 2016).

literature have reported a significant reduction in the risk of convulsions with magnesium sulphate (MgSO4) administration to women with severe pre-eclampsia (Edward et al. 2021; Kreepala et al. 2018). The multinational Collaborative Eclampsia Trial reported a 52% decrease of recurrent seizures among eclamptic women when treated with MgSO4 compared to other anticonvulsants (ACOG, 2020). This was similar to the World Health Organization’s 2011 recommendations, where magnesium sulphate was established as the anticonvulsant of choice for the prevention and treatment of severe pre-eclampsia and eclampsia (Imaralu & Jo, 2018).

The average serum magnesium (Mg) level in severe pre-eclampsia patients tend to increase substantially after administration of magnesium sulphate from 1.39 mmol/L to 4.90 mmol/L (Lumbanraja, Adnani, Dina, Lubis, & Hartono, 2020). Studies have reported that MgSO4 after administration tend to be tracible to both maternal and fetus.

MgSO4 is distributed in such a way that only 40% of plasma Mg becomes protein-bound while the rest, unbound Mg diffuses into diverse physiological compartments, including the bones, extracellular space. Furthermore, some Mg ions cross the placenta and fetal membranes reaching both the fetus and the amniotic fluid (Altraigey & Mostafaa, 2019; Darma, Nindrea, Idaman, Zaimy, & Irman, 2021; Das, Chaudhuri, Mondal, Mitra, Bandyopadhyay, et al., 2015; Nightingale, 2012).

The recommended serum Mg concentration for prevention of eclamptic convulsions is within the range of 1.8 to 3.0 mmol/L. However, close monitoring of Mg levels is important because of the potential consequences associated with deviations from this therapeutic window. Sub-therapeutic levels may result in the control of severe pre-eclampsia and prevention of eclampsia, leading to favorable obstetric outcomes. On the other hand, supra-therapeutic levels i.e. reaching concentrations above the recommended levels induces Mg toxicity. This toxicity poses potential risks, including severe conditions such as cardiac arrest which may cause death (Hassan, Elhhatim, Bakhit, & Shrif, 2014; J. O. Imaralu & Jo, 2018; Kreepala et al., 2018; Lumbanraja et al., 2020; Nightingale, 2012; Okusanya, Oladapo, Long, Lumbiganon, Carroli, Qureshi, & Duley, 2016; Omu, Al-Harmi, Vedi, et al., 2008). This narrow window of therapeutic serum magnesium range in managing severe pre-eclampsia calls for necessity of close monitoring to optimize the effectiveness of eclampsia treatment while minimizing the associated risks of both inadequate seizure control and magnesium toxicity.

Several factors have been reported to be determinant of therapeutic levels of Mg in managing severe preeclampsia. These factors include, body mass index (BMI), gestational age at the time of administration, altered renal function, altered liver function, diabetes mellitus, and the dosage of MgSO4 administered (Okusanya, Oladapo, Long, Lumbiganon, Carroli, Qureshi, & Duley, 2016; Titapant & Mingsuttiporn, 2020; Xavier, Johansson, Marcela, Galvão, et al., 2020). The body mass index (BMI) has been reported to be a critical determinant of the therapeutic levels of magnesium sulfate (MgSO4), this indicates the potential influence of body weight on magnesium distribution and metabolism. Patients with higher BMI requires adjustment in MgSO4 dosage so as to achieve recommended therapeutic levels which accounts for variations in the volume of distribution within the body (Choi et al. 2021). Also, gestational age at the time of MgSO4 administration is reported to determine the therapeutic levels of Mg, it is related to the dynamics of physiological changes due to pregnancy. Studies revealed that there is necessity of considering dosage adjustment with regard to gestation age because as pregnancy progress there is increment of maternal plasma volume and alteration in renal function that can directly impact the magnesium pharmacokinetics (Choi et al. 2021; Leetheeragul et al. 2020; Titapant and Mingsuttiporn 2020; Tudela and Alexander 2013).

Renal function is another factor affecting magnesium concentration in serum, because kidneys play a role in the retention and excretion of magnesium. So, abnormal kidney function can disrupt the normal clearance of magnesium, this will eventually lead to elevated magnesium levels and increased risk of toxicity. Understanding the status of renal function of the patient is very crucial in tailoring MgSO4 therapy to individual needs. Furthermore, it has been found that the presence of diabetes is also associated with altered magnesium metabolism. This adds another layer of complexity in managing severe pre-eclampsia, diabetic patients require special attention to ensure the achievement of therapeutic magnesium levels (Das, Chaudhuri, Mondal, Mitra, & Bandyopadhyay, 2015; Leetheeragul et al., 2020)

The administered dosage of MgSO4 is among modifiable factor that directly determines the serum concentration of Mg. Therefore, having the right choice in dosage, considering gestational age, BMI, renal function, and other individual factors, is crucial in avoiding both sub-therapeutic and supratherapeutic levels that could precipitate magnesium toxicity. For example, one systematic review reported the base line of serum magnesium to be <1.00 mmol/L with mean levels ranging from 0.74 to 0.85 mmol/L. After administering intravenous regime of 4g loading dose, serum magnesium levels raised sharply to about twice the baseline levels at half an hour being 1.48–1.70 mmol/L. At first, second, and fourth hours of the maintenance dose of 1g, the mean serum levels remained at a fairly constant level that was consistent with the values attained at first 30 minutes (Subedi, Bhansakarya, Shrestha, & Sharma, 2020).

The maintenance of serum therapeutic levels of magnesium sulfate is crucial in preventing undesirable obstetric outcomes. Subtherapeutic levels of magnesium sulfate lead into uncontrolled seizures which pose a significant risk of complications for both the maternal and the fetus. In cases where serum magnesium levels fall below the therapeutic range, the effectiveness of controlling seizure diminishes which result into prolonged or recurrent seizures which end up with maternal injury and compromise fetal well-being. On other hand, magnesium toxicity is reported to cause adverse neonatal outcomes, including hypotonia, bradycardia, birth asphyxia, the need admission to the neonatal intensive care unit (NICU) and interventions like intubation in the delivery room (Altraigey & Mostafaa, 2019; Das, Chaudhuri, Mondal, Mitra, Bandyopadhyay, et al., 2015; Duffy, Odibo, Roehl, MacOnes, & Cahill, 2012; Zarean & Tarjan, 2017).

According to hospital based unpublished data at Iringa Regional Referral Hospital (IRRH), severe preeclampsia is among the top ten disorders encountered in the antenatal ward. Also, MgSO4 is the medication of choice for treating severe pre-eclampsia. Despite the consistent use of MgSO4 in managing severe preeclampsia at the hospital, the occurrence of adverse obstetric outcomes remains notably significant. This highlights the necessity for conducting research to explore potential factors leading to unsatisfactory levels of adverse outcomes related to severe pre-eclampsia. Therefore, this study aimed to explore the potential factors associated with levels of serum Mg and their correlation with obstetric outcomes among severe preeclamptic women at IRRH.

## Materials and Methods

### Study design

This was a hospital-based analytical cross-sectional study design.

### Study duration

The study was conducted in six months spanning from November 2023 to April 2024.

### Study area

This study was conducted at IRRH in antenatal ward. The hospital receives women referred from the district hospitals surrounding Iringa town, private hospitals, health centres and dispensaries. Apart from receiving referrals from primary facilities, IRRH also accepts self-referral patients from home or traditional birth attendants. The Obstetrics and Gynaecology department at IRRH has four wards which includes gynaecological ward, antenatal ward, labour ward and postnatal ward. IRRH has capacity of 80 beds in obstetrics and gynaecology department. The department has four obstetricians/ gynaecologists, 2 medical doctors, 1 assistant medical officer, 51 nurses and obstetrics/gynaecology residents capable of managing obstetric emergencies. It also has a well-functioning neonatal intensive care unit. The hospital has two ICUs with 10 beds shared by all departments. Patients needing ICU care are transferred to the unit after interdepartmental consultation of the doctors in the obstetrics and gynaecology and those in the ICU. The hospital has 4 well equipped operating rooms.

Iringa Regional Referral Hospital receives approximately four patients with severe pre-eclampsia per week making total of 20 patients in a month. All patients with severe pre-eclampsia are managed by using MgSO_4_ to prevent progress into eclampsia.

### Study population

All pre-eclamptic women who were admitted at IRRH during the study period.

### Inclusion and exclusion criteria

#### Inclusion criteria

A woman with BP equal to or more than 160/110 mmHg and protein ++ in urine with one or more of the following symptoms; Headache, blurring vision, epigastric pain or reduced urine output who have not yet started MgSO_4_.

#### Exclusion criteria

A woman with BP equal to or more than 160/110 mmHg and protein ++ in urine with one or more of the following symptoms; Headache, blurring vision, epigastric pain or reduced urine output who have not yet started MgSO_4_ and is hemodynamic unstable and have other comorbidities like DM, epilepsy and sickle cell diseases.

### Sample size

The sample size was calculated using Leslie Kish formula (1965) for continuous data in single population study (Charan& Biswas, 2013).

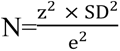

Where:

N = sample size

Z = is a standard normal variate (at 5% type1 error (p< (0.05) It is 1.96. e= absolute error which is 0.05 at 95% confidence interval

SD= standard deviation of serum Magnesium in pre eclamptic women as reported in previous study. Which was 0.28 mmol/L as reported by (Lumbanraja et al., 2020)

N= 120.47 approximately to 121

Therefore 121 women with severe pre-eclampsia during the study period were enrolled in this study.

### Sampling method

Study participants were selected by non-probability, convenient sampling technique where by women with severe pre-eclampsia admitted at IRRH during the study period and met the inclusion criteria were enrolled to the study until the sample size was reached.

### Data collection tools and procedures

#### Data collection tools

A comprehensive approach of data collection was achieved by using Swahili-translated structured questionnaire which was administered through interviews by the researcher. The questionnaire gathered information on various facets, including sociodemographic characteristics of participants, their obstetric history, gynecological history, and additional clinical details related to severe pre-eclampsia. The questionnaire captured information related to factors associated with pharmacologic serum levels of Mg. Additionally, the instrument included sections dedicated to record observed data derived from physical examinations, laboratory investigations, and obstetric outcomes.

#### Study procedure

The study team was composed of one principal investigator (PI) and one research assistant (RA) who approached the potential participants in antenatal ward and screened them for eligibility into the study. Women diagnosed with severe pre-eclampsia with gestation age of 28 weeks and above at IRRH during the study period were approached and introduced about the aim of the study. Only those who met inclusion criteria and voluntarily gave an informed consent to participate in this study were enrolled. Unique identification number was given and filled in the structured questionnaires.

Thorough history taking and physical examination was carried out by the PI and RA. Blood pressure (BP) was measured by using aneroid sphygmomanometer with an adult size cuff while the patient in a sitting up position; the arm being at the level of the heart. Those patients with BP equal to or more than 160/110 mmHg and protein ++ in urine with one or more of the following symptoms; Headache, blurring vision, epigastric pain or reduced urine output were recruited to the study, baseline serum Mg concentration were measured prior to initiation of loading dose of MgSO_4,_ and measured again at 4 hours and 12 hours after initiation of MgSO_4_.

Using a 5cc syringe, 4 millilitres of blood was taken from a vein at the cubital fossa of the non-dominant hand, placed in a red top tube (plain tube) and taken to the laboratory within 15 minutes. Samples were taken by PI or RA who are health personnel’s working in antenatal ward.

In the laboratory, the sample were left to settle for half an hour in room temperature to allow clotting. After settling it was centrifuged to obtain serum at 3000 rpm for 3mins using Centrifuge machine (Horizon Model 653V 230V DRUKER 20PL) with serial number: 160489AA028 manufactured in USA. The obtained serum was put to the fully automated biochemistry machine Archtech ci 4100 with serial number: C400360 manufactured by Abbot USA. Results for serum concentration of Magnesium, ASAT, ALAT and LDH were available within half an hour. These processes were done by a selected laboratory technician specialized in biochemistry as per IRRH standard laboratory protocol. Random blood glucose was measured and recorded to all women recruited for the study, Glucometer (Accu-Chek Active Model: GB SN: GB 30675507) was used during the study.

Using an RGZ-120 Health weighing scale, the patient’s weight in kilograms was determined and rounded to the closest decimal place. The participants were instructed to face the investigator and stand firmly on a scale. They were told to take off their shoes, dress simply, and set their belongings on a nearby table. We then looked at the scale’s display to find out how much body mass they had. Each person’s height in centimeters was measured using a stadiometer (RGZ-120 Health scale). A direct reading was obtained and converted to meters to one decimal place after the scale was adjusted to touch the top of the participant’s head. By dividing their weight in kilograms by the square of their height in meters, body mass index was calculated.

### Definition of Variables

#### Independent variables

In this study the independent variables were sociodemographic characteristics including mother’s age in numbers, residence as urban and rural, gestation age in weeks, marital status as married and not married, level of education as formal and informal education and maternal occupation as peasant, employed. Obstetric history included parity, history of severe pre-eclampsia, diabetes mellitus, premature delivery and IUFD.

Predictors of therapeutic serum magnesium levels including BMI which was categorized as underweight (<18 kg/m^2^), normal weight (18-24 kg/m^2^), overweight (25-30 kg/m^2^) and obese >30kg/m^2^). Also, liver function test will be measured during admission of the patients where by ASAT 0 t0 35U/L and ALT of 7 to 56U/l will be considered normal. Also, kidney function test will be addressed by checking serum creatinine levels where 0.6 to 1.1 mg/dl will be considered normal range.

#### Dependent variables

Primary outcome of our study is pharmacologic serum concentration of magnesium and secondary outcome was obstetric outcome.

Pharmacologic serum magnesium concentration will be categorized into two groups which are;

- Therapeutic level (1.8-3.0 Mmol/L) and
- Non-therapeutic level
- `o` Sub therapeutic (<1.8 Mmol/L)
- `o` Supratherapeutic(>3.0Mmpl/L)

Obstetrics outcome for both maternal and fetus were grouped into two groups which are

- Desirable/good outcomes
- Adverse outcomes Favourable/good obstetric outcome

Uneventful recovery to both foetus and mother.

Unfavourable maternal outcome

Unfavourable maternal outcome encompassed all eventful recovery and death; which are defined as follows

Primary postpartum hemorrhage-Blood loss of greater than or equal to 500mls accompanied by signs or symptoms of hypovolemia within 24 hours after the birth process.

Abruption placental diagnosed based on clinical signs and symptoms of painful vaginal bleeding, tense and tender abdomen, hypertonic uterus, decreased or absent fetal kick.

Acute kidney injury (AKI) as oliguria (urine output is less than 30mls/hour for 6 hours or less than 400mls/24hrs) non-responsive to fluid or diuretics.

Pulmonary oedema diagnosed based on clinical symptoms and sign like coughing up blood-stained sputum, difficulty in breathing and auscultation revealing fine basal crepitations.

Stroke diagnosed based on neurological findings of difficulty walking, loss of balance and coordination, difficulty in speaking or understanding others who are speaking, numbness or paralysis in the face, leg, or arm following the raised blood pressure during pregnancy.

HELLP syndrome diagnosed by clinical examination of developing yellowish colouration of the eyes and supported by laboratory findings of elevated serum level of ASAT, ALAT and LDH.

Maternal death Certified death of a participant who was recruited and on follow up.

#### Adverse foetal outcome

Prematurity which is delivery at a gestation age less than 37 completed weeks,

Low APGAR score a new born baby of a study participant who with an APGAR score of less than 7 in the fifth minute after birth.

Respiratory distress syndrome, diagnosed clinically by difficulty in breathing of the new born

NICU admission this means all new born who admitted to neonatal intensive care unit for further management.

Still birth this included all neonates with no signs of life at birth, APGAR score of zero.

If a participant develops one or more of these complications was considered to have adverse obstetric outcome and if developed none was considered to have no adverse outcome (favourable/Good outcome).

### Reliability and Validity

The questionnaire was first prepared in English and then translated in Swahili which later on translated back to English by another individual to check for consistency. The questionnaire was pre-tested in pilot study of 10 participant at IRRH and be discussed and modified with the aid of supervisors.

RA was medical personnel working in the department to ensure good management of the patients. RA was trained on how to collect and handle data and was tested several times until they reach performance of around 100% to ensure consistency and uniformity of data.

Laboratory processing of the samples were done according to standard operating procedures in an accredited laboratory by experienced laboratory technician. The machines were calibrated before running the sample by in order to ensure the results are valid. To make the results consistent only one laboratory scientist who has experience in clinical chemistry was selected for biochemical analysis.

### Data analysis

The data from completely filled questionnaires were entered in Microsoft Excel and cleaned. IBM SPSS program version 29 was used for data analysis in accordance to specific objectives. Descriptive statistics was used whereby continuous variables like mother’s age and gestation age were presented in median and categorical variables were analyzed and presented in percentage, frequencies in tables and graphs.

Inferential statistics was used to assess the factors associated with pharmacologic levels of serum magnesium among severe pre-eclamptic women administered with MgSO_4_ and correlation between pharmacologic levels of serum magnesium and obstetrics outcome. Chi square test and binomial logistic regression model was adopted to detect the association between the potential predictor variable and outcome variables. univariate logistic analysis was done to obtain crude odds ratio then the factors showing association were re-analyzed in Multivariable logistic analysis to obtain the adjusted odds ratios (removing the confounders) to determine the factors associated with magnesium sulphate concentration. 95% confidence interval was computed and P-value less than 0.05 was considered statistically significant.

### Ethical Clearance and Consideration

Ethical clearance was sought from the University of Dodoma (UDOM) Institutional Research Review Ethical Committee (IRREC). Permission to conduct study was sought from the hospital director of IRRH. To ensure confidentiality participants were coded with numbers instead of their real names. Documents were placed in a secure place which was only accessed by principal investigator and research assistants. Electronic data was be protected by password.

Written informed consent was obtained from participants before being enrolled in the study. Full elaboration of the study aims and benefit was done to the participants and they had chance to voluntary participate in the study. Participants were informed that they can withdraw from the study at any stage of the interview if they so desire and their withdrawal or decline in participating was not going to prevent them from being given equal care as other study participants.

## RESULTS

### Socio-Demographic and Clinical Characteristics of Women with Severe pre-eclampsia admitted for delivery at IRRH

The median age of the study participants was (29.0±10 IQR) years where 66 (54.5%) were in the age group of 25-34 years. 75 (62.0%) participants were residing in urban whereas 62(51.2%) had non or primary education. 83 (68.6%) were referred from lower facilities and 74 (61.2%) were multiparous (Table 1). Regarding the clinical features, 100(82.6) participants had headache, 65(53.7%) epigastric pain, 48(39.7%) reduced urine output, Protein in urine +++ were 43(35.5%) and protein ++ were 38(31.4%), 33 (27.3%) had reduced fetal movement and 15 (12.4%) with no fetal movement and abnormal heart rate were 23(19.0%) (Table 2)

### Pharmacologic Serum Concentration of Magnesium Among Women with Severe pre-eclampsia Before Administration of MgSO_4_

At admission before administration of loading dose of MgSO_4_ the Median serum concentration of Mg was (0.80 ±0.29 IQR) Mmol/L. 59 (48.8%) women had serum concentration within reference range of 0.75 - 1.05 Mmol/L. 53 (43.8%) women had low serum concentration below 0.75 Mmol/L and only 9 (7.4%) women had high serum concentration above 1.05 Mmol/L (figure 1)

**Figure 1:**
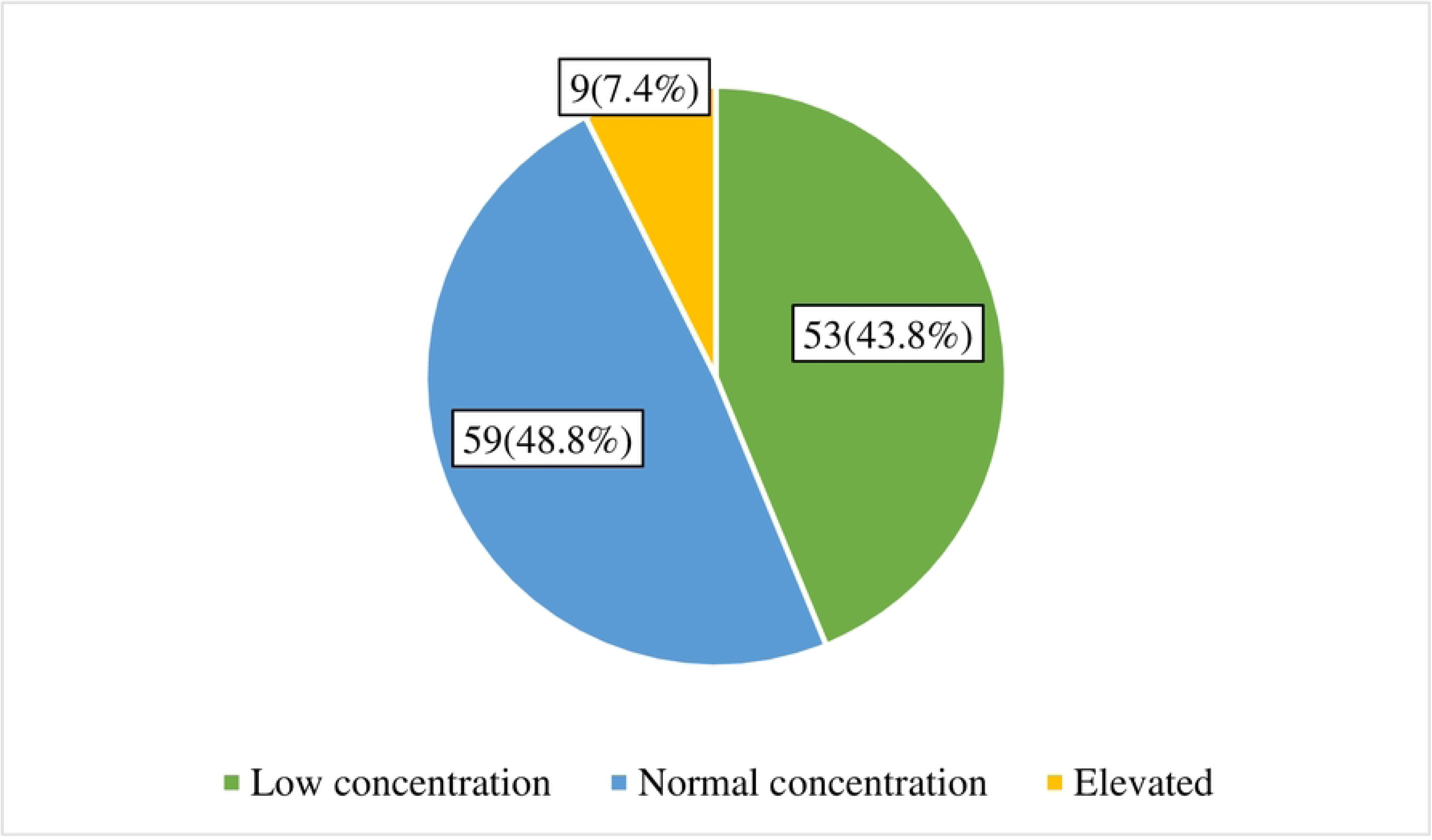
Pharmacologic serum concentration of Mg among women with severe pre­ eclampsia admitted at IRRH prior to administration ofMgS04 (N=121)

### Characteristic of serum concentration of Magnesium among women with severe pre-eclampsia admitted at IRRH for delivery at 4 and 12 hours post loading dose of MgSO_4_ administration

Serum Mg was measured at four hours and at 12 hours post loading dose of MgSO4 administration. Comparing concentrations at four hours versus at 12 hours it was observed that the proportion of women who attained therapeutic levels at four hours were 74(61.2%) and at twelve hours were 101(83.5%) (Figure 2). The median magnesium concentration at four hours was (1.9±1.02) Mmol/L whereas at 12 hours was (2.0±1.0) Mmol/L.

**Figure 2:**
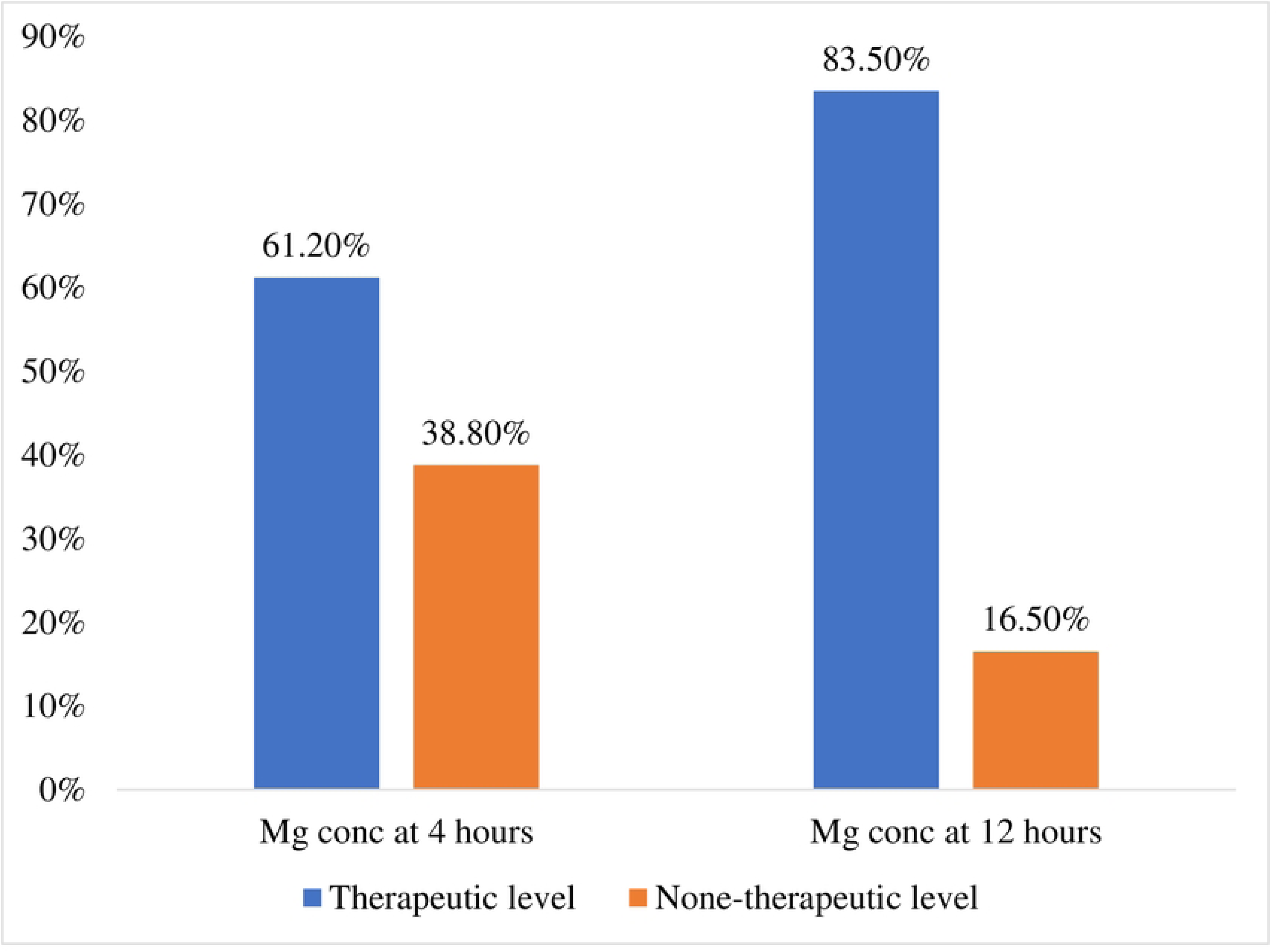
Characteristic of serum Mg concentration at 4 hours and 12 hours post loading dose of MgS04 (N=121)

### Factors Associated with Pharmacologic Serum Concentration of Magnesium Among Women with Severe pre-eclampsia admitted at IRRH for delivery

In chi square test, several factors were significantly associated with pharmacologic serum magnesium concentration among women with severe pre-eclampsia. Such factors included Residence (P value=0.049), Body mass index (0.002), creatinine levels (<0.001), serum Magnesium concentration at admission (<0.001), and proteinuria (<0.001) (table 3).

In multivariate regression, it was found that pregnant women with proteinuria +++ were seven times at increased risk of not attaining therapeutic serum Mg concentration. [P-value= 0.004, OR=7.13, 95% CI (1.89-26.86)]. Women who were Overweight were six-times at increased risk of not attaining therapeutic serum Mg concentration [P=0.002, OR= 5.52, 95% CI (1.83-16.68)]. Women with elevated creatinine levels were five times at risk of not attaining therapeutic Mg concentration [P= 0.012, OR=4.49, 95% CI (1.38-14.60)]. Women who presented with low serum Mg concentration at admission were nine times at risk of not attaining therapeutic serum Mg concentration [p-value<0.001, OR=9.17, 95% CI (3.07-27.37) (Table 4).

### Association Between Pharmacologic Serum Concentration of Magnesium and Corresponding Obstetric Outcomes Among Women with Severe pre-eclampsia Admitted for Delivery at IRRH

Maternal adverse outcomes were 63(52.1%) and fetal adverse outcomes were 98(80.2%) which were unbearable high (figure 3). This study assessed obstetric outcomes with regard to women with therapeutic and none therapeutic serum magnesium concentration. It was found that among women with non-tharapeutic serum magnesium; 14(29.8%) ended up having AKI, 18(38.3%) had PPH, 11(23.4%) had abruption placenta which was higher compared to those who attained therapeutic levels of magnesium sulpahate. However, there were no statistical significant difference among women with therapeutic and non therapeuticlevels except for AKI which had a p value of 0.048 (Table 5).

**Figure 3:**
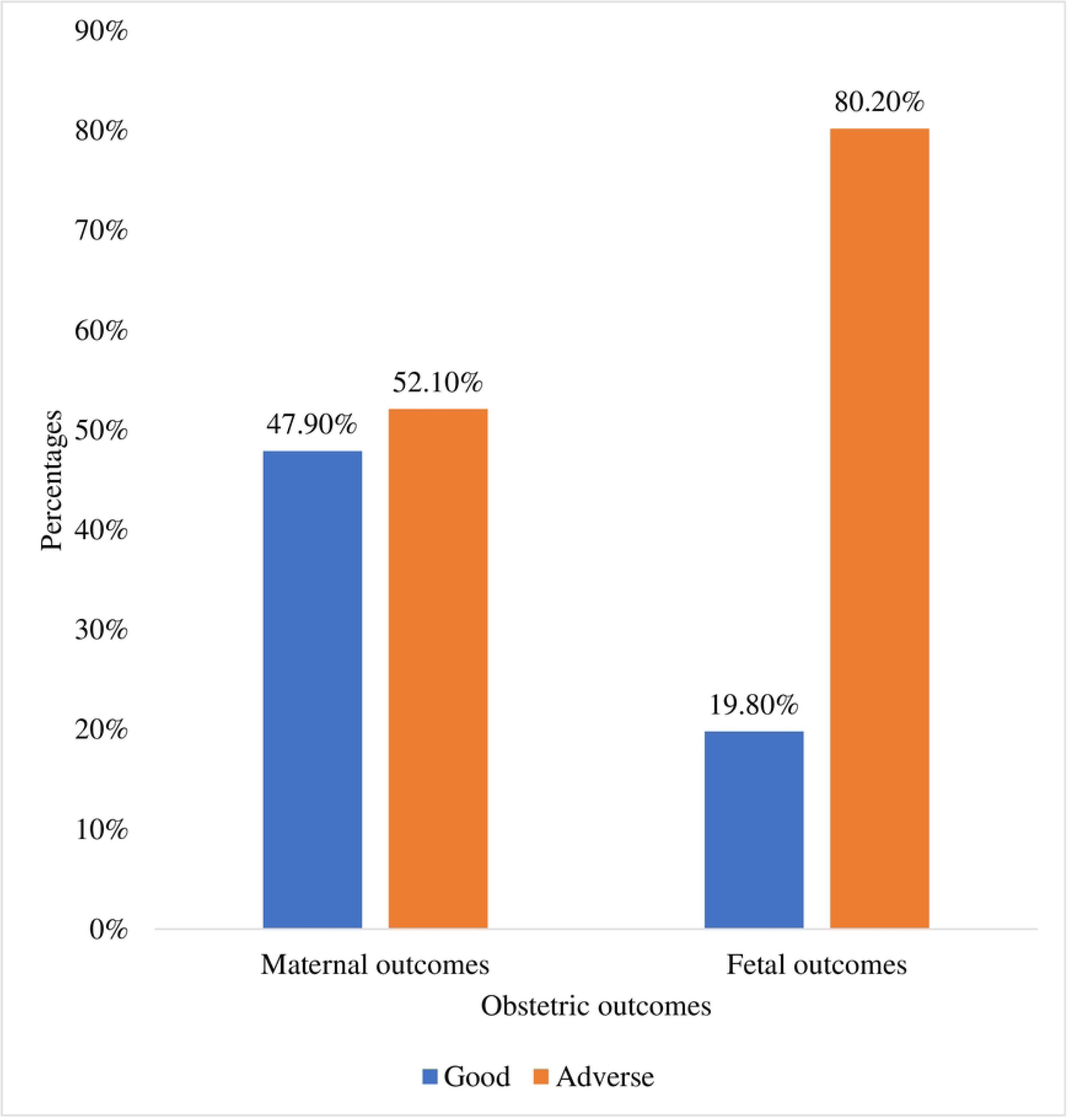
Overall obstetric outcomes among women with severe pre-eclampsia admitted fo delivery at IRRH.

## DISCUSSION

### Serum Concentration of Magnesium Among Women with Severe pre-eclampsia admitted for delivery at IRRH

The findings were similar to the study done in Brazil which reported the baseline Mg among severe pre-eclamptic women to be 0.77 mmol/L (1.87 mg/dL) which is within normal physiological range (Xavier, Johansson, Marcela, Galvão, et al., 2020). Another study has showed that the mean serum Mg level in the severe preeclampsia group was significantly lower than that in the control group, with levels of 0.7 mmol/L in the severe preeclampsia group compared to 0.77 mmol/L in the control group (p = 0.004)(Lumbanraja et al., 2020).

A study done in China reported the baseline Mg level to be 0.76 mmol/L which is within normal physiologic range as in our study (Deng et al., 2024), similarly to the meta-analysis which included eight studies, the findings revealed that serum concentration of Mg is consistently less than 1mmol/L (Okusanya, Oladapo, Long, Lumbiganon, Carroli, Qureshi, Duley, et al., 2016). The serum concentration of Mg was noted to be lower in significant number of pre eclamptic in this study. This was reported in several other studies showing that Mg tend to be lower among women with pre-eclampsia than health women (Adekanle et al., 2014). I Nigeria the study have reported the concentration of 0.69 mmol/L which was significantly lower compared to normotensive pregnant women (Igberase, Ebeigbe, Okonta, Okpere, & Gharoro, 2007).

Pregnant women may experience raised Mg demand due to the needs of the growing fetus and placenta which exacerbating any preexisting deficiencies. Furthermore, conditions like increased renal clearance during pregnancy, dietary insufficiencies specific to pregnancy, and the stress of pregnancy can further deplete Mg levels (Joob & Wiwanitkit, 2019; Nagaria, Mitra, & Banjare, 2017; Yousef, Merghani, Abdulla, & Binni, 2022).

### Characteristic of Serum Concentration of Magnesium Among Women with Severe pre-eclampsia After MgSO_4_ Administration

The findings show that 61.2% of patients attained therapeutic levels at four hours. The proportion of patients who were under therapeutic level of serum magnesium was higher at 12 hours 83.5%. The median magnesium concentration at four hours was 1.9 with IQR of 1.02 Mmol/L whereas at 12 hours was 2.0 with IQR of 1.0 Mmol/L. Other previous studies have reported similar findings showing that majority of women attained therapeutic levels after administration of magnesium sulphate.

A meta-analysis of multiple studies revealed that following the loading dose of magnesium sulfate (4 g), serum magnesium levels rose sharply to approximately twice the baseline levels within half an hour, reaching between 1.48 and 1.70 mmol/L. These levels remained fairly constant at 1, 2, and 4 hours of the maintenance dose, maintaining the same range even at 8, 12, and 24 hours, without ever exceeding a mean serum concentration of 2.00 mmol/L. This consistency aligns with a steady-state level of 1.64 mmol/L and an average concentration of 1.70 mmol/L reported in other previous studies. In addition, some studies indicated that the peak serum concentration was achieved within the first half-hour of treatment initiation. These findings underscore the rapid attainment and maintenance of therapeutic magnesium levels with this dosing regimen (Jonsdotter, Rocha-Ferreira, Hagberg, & Carlsson, 2022; Li et al., 2020; Okusanya, Oladapo, Long, Lumbiganon, Carroli, Qureshi, Duley, et al., 2016).

In Chinese patients large proportion of the study participants did not attain the therapeutic level of magnesium sulphate accounting 55.9% after administration of the loading dose (Li et al., 2020), this proportion was very high compared to what was found in this study which was 38.8 women who didn’t attain therapeutic level at four hours post magnesium administration. Another study revealed that before the administration of MgSO4, the average magnesium level in preeclampsia patients was 1.39 mEq/L (SD ± 0.28), the concentration rose dramatically to 4.90 mEq/L (SD ± 0.37) following MgSO4 administration (Lumbanraja et al., 2020).

The study done in Sweden have revealed that None of the women experienced magnesium toxicity but about 72% of the women showed serum magnesium levels within the therapeutic range (2.0– 3.5 mmol/L) and no adverse events were observed during the infusion. The serum magnesium levels in the mothers were steadily same until 24 hr after delivery when it declined to pre-bolus levels (Jonsdotter et al., 2022). The proportion of women reached therapeutic levels was slightly higher to our study which showed 61.2%. the main difference in these studies is that in the study done in Sweden used loading dose of 6g while in this study it was 4g. another difference was seen in reference range of therapeutic levels which in our study was 1.8-3.0 Mmol/l which was different from that of Sweden.

This study was in line with previous studies which in demonstrating distinct pattern in serum magnesium levels following the administration of magnesium sulfate in women with preeclampsia or preterm labor. The findings showed that serum magnesium levels rose sharply reaching approximately twice the baseline levels. These elevated levels remained relatively constant at subsequent time points that is 4, and 12hours. This consistency in therapeutic levels underscores the effectiveness of MgSO_4_ in management of severe pre-eclampsia.

### Factors Associated with Pharmacologic Serum Concentration of Magnesium Among Women with Severe pre-eclampsia admitted for delivery at IRRH

In this study overweight was found to be significant predictor of subtherapeutic levels of serum magnesium at four hours. The findings showed that women who are overweight was about six-fold risk of attaining none-therapeutic magnesium concentration. This was similar to the study done in Texas which reported that greater BMI was independent predictor of subtherapeutic (Tudela & Alexander, 2013). Another study has reported that body weight statistically influenced the clearance of MgSO_4_, indicating that patients with different body weights exhibit varying rates of drug clearance. This is a common finding in pharmacokinetics, as overweight individuals often have higher clearance rates due to their greater blood volume and enhanced kidney function (Xavier, Johansson, Marcela, & Galvão, 2020). In a study done in Thailand, it was found that body weight impacts serum magnesium levels in such a way that women with a BMI over 25 kg/m² are more likely to have subtherapeutic serum magnesium. Furthermore, the study found no cases of seizures in patients with subtherapeutic magnesium levels in either BMI group, and no instances of supratherapeutic magnesium levels were observed with a magnesium sulfate infusion rate of 1 gram per hour (Jaisamut & Kitiyodom, 2017). Joob and Wiwanitkit (2019) have reported that BMI is one among on admission factors that can affect the attainment of therapeutic levels of serum magnesium among patients with severe pre-eclampsia.

A study conducted in China found that women with a pre-pregnancy BMI greater than 25 kg/m^2^ had a 56% higher risk of subtherapeutic serum magnesium levels compared to those with a normal BMI. In other hand, women with a pre-pregnancy BMI less than 18 kg/m² had an 80% reduced risk of low magnesium levels. Similar to our study being overweight or obese is significant risk of subtherapeutic serum magnesium levels, however the study done in China evaluated pre-pregnancy BMI while this study evaluated the index BMI (Titapant & Mingsuttiporn, 2020).

Several factors can be the reason behind the fact that overweight affects the attainment of therapeutic serum levels. This can be explained by the reason that overweight and obese individuals tend to have a higher volume of distribution for drugs like magnesium sulfate. Drugs tend to disperse more extensively into body tissues which lead to a dilution of the drug concentration in the bloodstream (Choi et al., 2021). Another possible fact is explained by blood volume being high in overweight individuals enhancing kidney function, this fact is often linked with increased clearance rates of medications including magnesium sulphate leading into subtherapeutic levels (Okusanya, Oladapo, Long, Lumbiganon, Carroli, Qureshi, Duley, et al., 2016). Several scholars have reported the variation of metabolic processes with body weight. Obesity alters liver enzyme activity along with other metabolic pathways which impact how the drug is processed and eliminated. Standard dosing regimens may not adequately account for the increased body mass in overweight patients which lead to insufficient drug doses affecting the attainment of therapeutic levels of magnesium sulphate. Furthermore, inflammatory and hormonal factors may also play a role as overweight individuals often have altered levels of inflammatory markers and hormones that can affect drug absorption, distribution, metabolism, and excretion (Muganyizi & Shagdara, 2011; Okunade, Oluwole, & Adegbesan-Omilabu, 2014; Okusanya, Oladapo, Long, Lumbiganon, Carroli, Qureshi, Duley, et al., 2016; Tudela & Alexander, 2013).

This study found that pre-eclamptic women with significant proteinuria (+++) are seven times more likely to attain non-therapeutic magnesium concentrations compared to those without such severe proteinuria. Also, women with elevated creatinine levels are about five times more likely to attain non-therapeutic magnesium levels. This was similar to the study done in China which showed the elevated creatinine level was independent predictor of subtherapeutic levels of serum magnesium (Li et al., 2020). This high level of protein in the urine indicates substantial kidney dysfunction, as the kidneys are unable to retain essential proteins and this elevated creatinine signifies impaired kidney function, as it reflects the reduced ability of kidney to filter waste products from the blood. Both proteinuria and elevated creatinine levels are markers of compromised kidney function, which can affect the body’s ability to maintain therapeutic drug levels due to altered drug clearance and distribution. Serum creatinine was a marker of kidney function which influenced the volume of distribution of MgSO_4_ (Igberase et al., 2007; Li et al., 2020; Pascoal et al., 2019). Another study done in Thailand have reported renal clearance to be the significant factor for subtherapeutic serum magnesium among patients with severe pre-eclampsia, however the study used BUN and uric acid to assess the renal function while this study used serum creatinine (Leetheeragul et al., 2020).

In pre-eclamptic women, elevated serum creatinine levels and significant proteinuria indicate kidney dysfunction. This dysfunction directly impacts the pharmacokinetics of magnesium sulfate as it results into reduced clearance and altered distribution of magnesium, causing the drug to accumulate in tissues rather than maintaining therapeutic levels in the bloodstream (Westermann & Zoysa, 2022). In addition, kidney dysfunction results in fluid retention, further diluting serum magnesium concentrations (Li et al., 2020).

Furthermore, this study found that pre-eclamptic women who presented with low serum magnesium at admission were about nine-fold at risk of sub-therapeutic serum magnesium after treatment with magnesium sulphate. Studies have shown that the effectiveness of magnesium sulfate depends on the body’s initial magnesium status. Women with already low serum magnesium may have a diminished response to treatment due to a larger deficit that the initial doses cannot adequately correct. The study done in Nigeria have reported that low serum magnesium level before or during treatment with magnesium sulphate is a risk factor for convulsion (Padda et al., 2021b).

Low level of serum magnesium is associated with likelihood of attaining subtherapeutic levels as studies have reported that women who received the 4-gram dose had lower serum magnesium levels compared to those who received the 6-gram dose and were less likely to achieve therapeutic magnesium levels. Even after adjusting with other risk factors like BMI and renal clearance it was found that women who received the 6-gram dose were more than twice as likely to achieve therapeutic magnesium levels compared to those who received the 4-gram dose (Westermann & Zoysa, 2022). Some studies have recommended the prophylactic management using magnesium supplementation to prevent eclampsia and hypomagnesemia among pregnant women. The supplementation can raise the serum levels of magnesium which can enhance the effectiveness of magnesium sulphate in treating severe pre-eclampsia (Leetheeragul et al., 2020).

### Association Between Pharmacologic Serum Concentration of Magnesium and Corresponding Obstetric Outcomes Among Women with Severe pre-eclampsia admitted for delivery at IRRH

The overall adverse maternal and fetal outcomes was found to be high in this study making up 52.1% and 80.2% respectively. The most encountered adverse outcomes included AKI 20.7%, PPH 29.8. Fetal outcomes included death 16.5%, NICU admission 81.8%, low Agar score 63.6% and prematurity 80.2%. Other studies have shown similar pattern of adverse outcomes including renal damage in 8.7%, preterm birth 55.2%. The perinatal mortality rate was 27 per 1,000 (Omu, Al-Harmi, Al-Ragum, et al., 2008). The prevalence of adverse maternal outcomes in ethiopia was 80.2% which was higher compared to 52.1% reported in this study (Godana, 2021). Several studies have reported high prevalence of adverse outcomes including PPH, AKI, need for blood transfussion, retinal detachment, liver dysfunction and motality however in this study there was no maternal death (Kobina et al., 2014; Mirkovic et al., 2020; Syoum et al., 2022)

Preeclampsia significantly increases the risk of several adverse obstetric outcomes, including postpartum hemorrhage (PPH), acute kidney injury (AKI), prematurity, and other complications. This can be explained by several facts including coagulation abnormalities and uterine atony which increase the risk of PPH. It also affects renal perfusion and causes proteinuria which contribute to AKI. In the efforts of mitigating maternal and fetal risks, early delivery is often necessary which lead to prematurity. Poor placental blood flow can cause intrauterine growth restriction (IUGR) and placental abruption, while severe forms like HELLP syndrome and progression to eclampsia can result in significant maternal and fetal morbidity and mortality (Darma et al., 2021; Duffy et al., 2012; Foumsou et al., 2022; Kassie et al., 2014).

After adjusting pharmacologic magnesium serum concentration with other factors in multivariate logistic regression it was found that women with sub-therapeutic magnesium serum concentration were at risk of having adverse maternal outcomes about 23-fold compared to those who attained therapeutic levels. Other factors were Epigastric pain which was three-fold at high risk of adverse maternal outcomes compared to their counterpart. Sub-therapeutic magnesium levels significantly increase the risk of adverse maternal outcomes due to magnesium’s critical role in numerous physiological processes, including muscle and nerve function, blood pressure regulation, and preventing seizures in preeclampsia. Inadequate magnesium levels can lead to uncontrolled hypertension, seizures, and poor muscle contractility, contributing to severe complications such as eclampsia, postpartum hemorrhage, and acute kidney injury(Padda et al., 2021a). Furthermore, epigastric pain is often a sign of severe preeclampsia or HELLP syndrome, indicating liver involvement and increased risk of maternal morbidity (Mirkovic et al., 2020).

Achieving therapeutic magnesium levels post-administration of magnesium sulfate is crucial for preventing adverse obstetric outcomes in preeclamptic women. A significant number of women do not attain therapeutic levels due to factors such as renal function impairment, and patients body mass index. Monitoring serum magnesium levels is essential because sub-therapeutic levels can fail to prevent seizures and other severe complications associated with preeclampsia. For instance, insufficient magnesium can lead to uncontrolled hypertension and increased neuromuscular excitability, raising the risk of eclamptic seizures. Moreover, magnesium acts as a vasodilator, helping to lower blood pressure and improve placental blood flow, thereby reducing the risk of placental abruption and fetal growth restriction. Studies have shown that magnesium sulfate effectively reduces the risk of eclampsia by approximately 58% and maternal death by 45% when therapeutic levels are maintained (J. Imaralu, Olaleye, Badejoko, Loto, & Ogunniyi, 2015; Padda et al., 2021a). Additionally, proper magnesium levels can decrease the incidence of HELLP syndrome, which is characterized by hemolysis, elevated liver enzymes, and low platelet count, further preventing severe maternal morbidity. Therefore, regular monitoring and timely adjustment of magnesium sulfate dosage are vital in ensuring optimal maternal and fetal outcomes in preeclamptic women (J. Imaralu et al., 2015; Leetheeragul et al., 2020; Li et al., 2020; Titapant & Mingsuttiporn, 2020).

## Conclusion

Significant number of women with severe pre-eclampsia had low serum magnesium concentration at admission prior to administration of loading dose of MgSO_4_. The number of women who attained subtherapeutic levels of serum Mg at twelve hours post loading dose was low compared to four hours post loading dose of MgSO_4_. The median serum concentration levels of magnesium were higher at 12 hours compared to that measured at four hours. Women with high BMI are at risk of not attaining therapeutic serum concentration of Magnesium which expose them to unfavorable obstetric outcomes. Renal function which can be noted by using serum creatinine and presence of protein in urine is an independent predictor of sub-therapeutic levels among women with severe pre-eclampsia managed with MgSO_4_. Subtherapeutic serum concentration of Magnesium is associated with unfavorable obstetric outcomes. The most common obstetric outcomes were post-partum hemorrhage, acute kidney injuries, and stroke. There was no maternal death in this study, however fetal death was encountered in some cases mainly due to prematurity.

## Data Availability

All relevant data are within the manuscript and its Supporting Information files.

## Dissemination of the findings

Findings will be presented at the department of Obstetrics and Gynecology at the University of Dodoma. The study report will be compiled and submitted to the University library and Iringa Regional Referral Hospital and manuscript will be prepared for peer review and journal publication.

## Authors’ contributions

Conceptualization, Data curation, Formal analysis, Investigations, Methodology, Resources, Writing – original draft, Writing – review & editing: Joshua Stephen Kileo.

Conceptualization, Methodology, Project administration, Resources, Supervision, Writing – review & editing: Shubi Matovelo, Gabriel Kitinusa.

## Acknowledgements

I thank almighty God for this opportunity to pursue master of medicine in obstetrics and gynecology. Special thanks to the department of Obstetrics and gynecology at the University of Dodoma for the support and guidance throughout the training period. To my supervisors Dr. Shubi Matovelo and Dr. Gabriel Kitinusa who have been not only mentors but also an inspiration toward hard work and commitment.

## Supporting information

**S1 Table: Socio-demographic characteristics of women with severe preeclampsia admitted for delivery at IRRH (N=121)**

**S2 Table: Clinical characteristics of women with severe pre-eclampsia of women with severe pre-eclampsia admitted for delivery at IRRH (N=121)**

**S3 Table: Chi square test of factors associated with pharmacologic serum concentration of Magnesium among women with severe pre-eclampsia admitted at IRRH for delivery (N=121)**

**S4 Table: Univariate and Multivariate logistic regression of factors associated with pharmacologic serum concentration of Magnesium among women with severe pre-eclampsia admitted at IRRH for delivery (N= 121)**

**S4 Table: Association between pharmacologic serum concentration of Magnesium and corresponding obstetric outcomes among women with severe pre-eclampsia admitted for delivery at IRRH (N=121)**

**S1 Figure: Pharmacologic serum concentration of Mg among women with severe pre-eclampsia admitted at IRRH prior to administration of MgSO4 (N=121)**

**S2 Figure: Figure 2: Characteristic of serum Mg concentration at 4 hours and 12 hours post loading dose of MgSO4 (N=121)**

**S3 Figure: Figure 3: Overall obstetric outcomes among women with severe pre-eclampsia admitted for delivery at IRRH.**

